# Determinants of Prevalence and Stages of Undiagnosed Hypertension

**DOI:** 10.1101/2024.05.21.24307708

**Authors:** Fiorella E. Zuzunaga-Montoya, Luisa Erika Milagros Vásquez-Romero, Joan A. Loayza-Castro, Enrique Vigil-Ventura, Carmen Inés Gutierrez De Carrillo, Víctor Juan Vera-Ponce

**Author notes:** Correspondence: Víctor Juan Vera-Ponce. Luisa Erika Milagros Vásquez Romero. Carmen Inés Gutierrez De Carrillo.

## Abstract

**Introduction:** Hypertension (HTN) has been classified into stages according to elevated blood pressure levels. It has been detailed that higher levels correlate with an increased risk of long-term complications.

**Objective:** To determine the prevalence and trends of different hypertension stages among Peruvians and to identify factors associated with these stages.

**Methods:** Data from the Demographic and Family Health Survey from 2014 to 2022 were analyzed. Blood pressure levels were categorized into normotension, high blood pressure (HBP), and HTN stages I, II, and III. The study included individuals aged 18 years and older, excluding those who had not had their blood pressure measured or had known histories of hypertension.

**Results:** The prevalences of HBP, HTN stage I, II, and III were 14.73%, 9.64%, 2.16%, and 0.64%, respectively. Factors such as gender, age, wealth index, alcohol consumption, daily smoking, and health conditions like obesity and diabetes significantly influenced the prevalence of the various stages.

**Conclusions:** The prevalence of HTN varies significantly depending on the stage. These findings underscore the necessity for public health strategies that are personalized and differentiated according to the specific stage of HTN to enhance effectiveness.

## Introduction

Hypertension (HTN) is traditionally defined by elevated systolic (SBP) and diastolic blood pressure (DBP) values ^(1)^. Globally, the prevalence of HTN is rising, representing an emerging public health crisis with profound implications for the development of cardiovascular diseases and global mortality ^(2)^. According to the World Health Organization (WHO), over one billion people worldwide live with HTN, constituting approximately 21% of the adult population ^(3)^.In Peru, the situation mirrors this global trend, with a prevalence of 20.76% ^(4)^

This condition has been classified into various stages according to elevated blood pressure levels. Higher levels entail an increased risk of long-term complications ^(5)^. Consequently, while numerous studies have explored the factors associated with the presence of HTN in general, few have delved into which specific patient characteristics predispose individuals to higher stages of hypertension ^(6)^.

Therefore, the current study focuses on the following objectives: 1) to determine the prevalence and trends of different HTN stages among Peruvian patients and 2) to identify factors associated with HTN, using data from a national survey conducted over nine years.

## Methods

### Design

A cross-sectional and analytical analysis of secondary data from the Demographic and Family Health Survey (ENDES), provided by the National Institute of Statistics and Informatics (INEI), was conducted. The evaluated years were from 2014 to 2022 ^(7)^. This study adhered to the STROBE (Strengthening the Reporting of Observational Studies in Epidemiology) guidelines to ensure high quality and transparency in reporting observational studies^(8)^.

### Population, Eligibility Criteria, and Sample

The ENDES focused on a representative sample of the Peruvian population, which included individuals aged 15 to 99 residing in both urban and rural areas of the country’s 24 departments. It utilized a probabilistic, stratified, and two-stage sampling design, differentiated for urban and rural areas, whose details are specified in the ENDES’s technical reports.

For the present study, individuals aged 18 years and older were selected for analysis, following standard definitions of arterial hypertension for this age group—participants with no blood pressure measured or those with known hypertension were excluded. Additionally, to ensure the inclusion of only valid blood pressure measurements for the analysis, specific thresholds were established: systolic blood pressure (SBP) had to be between 70 and 270 mmHg, and diastolic blood pressure (DBP) between 50 and 150 mmHg ^(9)^. Measurements outside these plausibility ranges were excluded from the study.

### Variables and Measurement

The main variables were as follows:

1. Normotension: Individuals with an SBP less than 130 mmHg and a DBP less than 80 mmHg.
2. High Blood Pressure (HBP): Those with an SBP of 120-139 mmHg or an 80-89 mmHg DBP.
3. Stage I Hypertension: Individuals with an SBP between 140 and 159 mmHg or a DBP between 90 and 99 mmHg.
4. Stage II Hypertension: Those with an SBP between 160 and 179 mmHg or a DBP between 100 and 109 mmHg.
5. Stage III Hypertension: Defined by an SBP of 180 mmHg or more or a DBP of 110 mmHg or more.

The variables considered in this analysis include sex (female and male), age grouped by ranges (18-35 years, 36-59 years, and 60 years and older), geographical location (Metropolitan Lima, other coastal areas, Highland, and Jungle), educational level (none, primary, secondary, and higher education), socioeconomic status (very poor/poor, middle, and rich/very rich), and type of residential area (urban or rural). Health behaviors such as daily smoking (yes or no) and alcohol consumption (never/non-excessive vs. excessive) were also examined. Nutritional status was assessed using the body mass index (BMI), classified as normal weight (BMI ≤ 24.99 kg/m^2^), overweight (BMI 25 to 29.99 kg/m^2^), and obesity (BMI ≥ 30 kg/m^2^), as well as the presence of abdominal obesity, defined as a waist circumference ≥ 102 cm in men and ≥ 88 cm in women. The presence of diabetes mellitus was evaluated through self-report (yes or no), and the altitude of the residence was classified into four categories: 0 to 499 m, 500 to 1499 m, 1500 to 2999 m, and 3000 m or more.

### Procedures

To follow a standardized method, an OMRON digital sphygmomanometer, model HEM-713, was used to measure SBP and DBP using the oscillometric blood pressure technique. Due to possible differences in arm circumference among participants, two cuffs of different sizes were employed according to individuals’ needs: one for standard-sized arms measuring 220-320 mm and another adaptable for larger arms with circumferences ranging from 330-430 mm.

Participants’ measurements took place under controlled conditions where they could relax; seated at rest with their right hand elevated to heart level when both forearms rested on a flat surface extending forwards without any other body part touching it. Participants were not supported by pillows underneath their forehand. The first BP value was taken after five minutes during the resting phase, followed by a second measurement two minutes later to achieve stabilization before entering into situations affecting cardiovascular conditions. Finally, a third measurement was taken just a few seconds ahead to determine proper values based on the overall tendency reflecting all potential influencable events impacting each distinct case presented within the group sample. The purpose was to minimize momentary variation, leading to higher reliability estimates associated with one’s baseline arterial blood pressure. Informed consent: Participants read and signed a consent form before participating in the study.

### Statistical Analysis

This study used the statistical software R version 4.03 to conduct all relevant statistical analyses. Initially, a descriptive analysis of all variables was performed to provide an overview of the characteristics of the studied population. This analysis included the distribution of frequencies, means, and standard deviations for quantitative variables and proportions for categorical variables, facilitating an initial understanding of the dataset.

A multinomial logistic regression model was implemented to analyze the factors associated with different stages of arterial hypertension. This approach was chosen due to the polytomous nature of the dependent variable, which categorizes hypertension into several stages. The model allowed for the estimation of adjusted odds ratios (aOR) for each hypertension stage compared to the normotensive group, including 95% confidence intervals (CI 95%) for each estimate. The model was adjusted for various demographic and behavioral covariates previously identified as relevant.

Finally, trend graphs were generated to visualize the results, showing the evolution of different stages of HBP and HTN from 2014 to 2022.

### Ethical Considerations

This study utilized publicly accessible and cost-free data, which were anonymized to protect the participants’ privacy by removing any information that could personally identify them, thus mitigating potential ethical risks. Additionally, the database, which is publicly available for review and follow-up, provides open access to the collected data without personal identifiers and direct interaction with human subjects^(10)^. Due to these conditions, it was considered unnecessary to submit this study for review by an ethics committee.

## Results

In the study involving 231,834 participants, the sex distribution was balanced, and most individuals were concentrated in the age range of 20 to 44 years (62.29%). A high percentage of the sample (76.28%) resided in urban areas, with a third specifically in Metropolitan Lima (33.76%). The study revealed that 41.28% of the participants belonged to the lowest wealth stratum. Regarding lifestyle behaviors, only 1.70% of the participants reported being active smokers, and 2.76% consumed alcohol excessively. Most participants (72.84%) presented normotension, while cases of stage I hypertension were the most prevalent among the different stages of hypertension, affecting 9.64% of the population. The rest of the characteristics can be seen in Table 1.

**Table 1.** Demographic and anthropometric characteristics of participants.

| **Characteristic** | **n = 231,834** |
| --- | --- |
| **Sex** |  |
| Female | 117,292 (50.59%) |
| Male | 114,543 (49.41%) |
| **Group age** |  |
| 20 - 44 years | 144,420 (62.29%) |
| 45 - 59 years | 53,746 (23.18%) |
| 60 to more | 33,668 (14.52%) |
| **Educational Level** |  |
| No Level | 458 (0.25%) |
| Primary | 42,180 (22.94%) |
| Secondary | 77,386 (42.09%) |
| Superior | 63,826 (34.72%) |
| **Natural region** |  |
| Metropolitan Lima | 78,268 (33.76%) |
| Resy of coast | 59,278 (25.57%) |
| Mountain Range | 63,888 (27.56%) |
| Jungle | 30,401 (13.11%) |
| **Wealth index** |  |
| The poorest/Poor | 95,698 (41.28%) |
| Medium | 47,170 (20.35%) |
| Rich/Richest | 88,967 (38.38%) |
| **Area of residence** |  |
| Rural | 176,848 (76.28%) |
| Urbano | 54,986 (23.72%) |
| **Smoker Status** |  |
| No | 227,892 (98.30%) |
| Yes | 3,942 (1.70%) |
| **Alcohol consumption** |  |
| Never or Non-excessive alcohol consumption | 225,442 (97.24%) |
| Excessive alcohol consumption | 6,392 (2.76%) |
| **Status Nutritional** |  |
| Normal weight | 79,887 (34.47%) |
| Overweight | 96,687 (41.71%) |
| Obesity | 55,217 (23.82%) |
| **Abdominal Obesity** |  |
| No | 72,597 (57.01%) |
| Yes | 54,753 (42.99%) |
| **Diabetes** |  |
| No | 225,349 (97.25%) |
| Yes | 6,365 (2.75%) |
| **Altitude** |  |
| 0 to 1499 | 164,092 (70.80%) |
| 1500 to 2499 | 15,736 (6.79%) |
| 2500 to 3499 | 32,726 (14.12%) |
| 3500 or more | 19,226 (8.29%) |
| **Blood pressure levels** |  |
| Normotension | 168,862 (72.84%) |
| HBP | 34,141 (14.73%) |
| HTN stage I | 22,338 (9.64%) |
| HTN stage II | 4,999 (2.16%) |
| HTN stage III | 1,493 (0.64%) |

### Trends in Different Stages of Arterial Hypertension

Figure 1 illustrates the prevalence trends of various HTN stages and HBP from 2014 to 2022. There is a general increase in the prevalence of HBP, rising from 14.76% in 2014 to 15.15% in 2022, with a peak of 16.58% in 2021. Stage I HTN decreased, starting at 9.72% in 2014 and declining to 10.74% in 2022. On the other hand, cases of stage II and stage III HTN, although less common, showed minor fluctuations, with a slight decrease in 2022 compared to the previous year. Stage II HTN hovered around 2% throughout the period, while stage III HTN remained below 1%, indicating a relatively stable but low prevalence.

**Figure 1.**
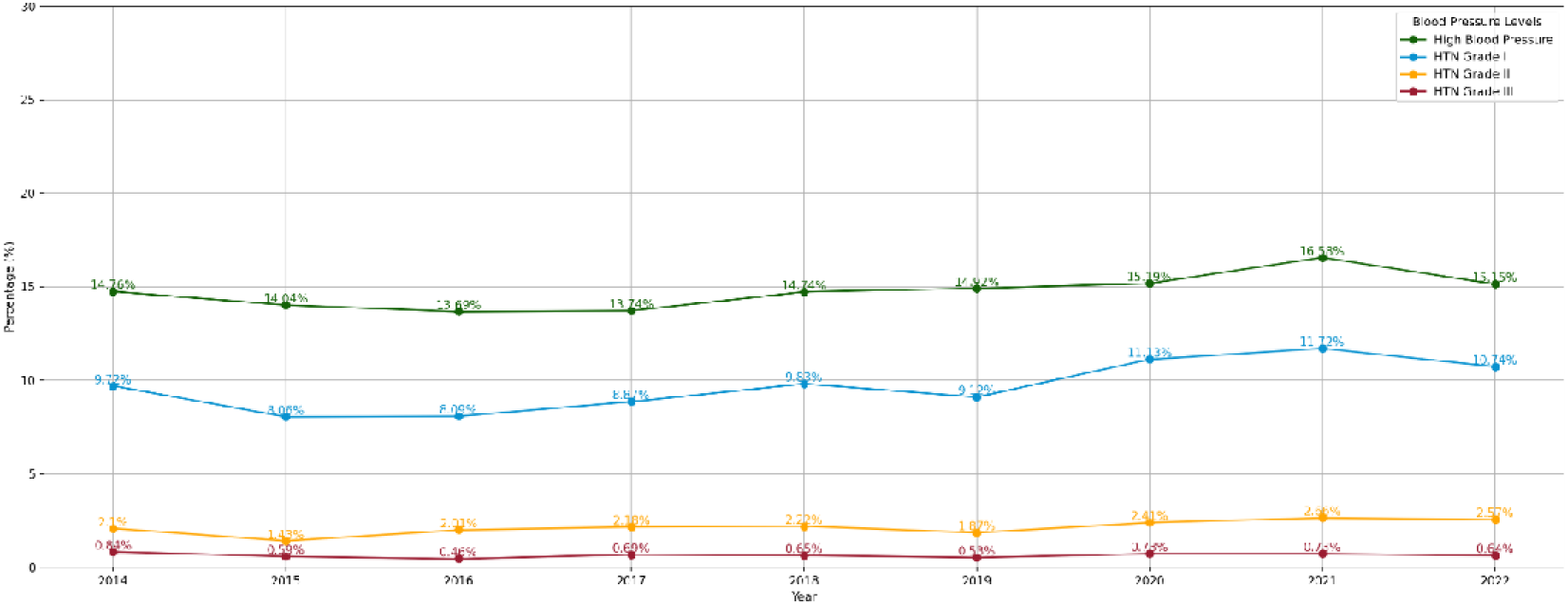
Trends in different stages of arterial hypertension.

### Multivariable analysis of factors associated with different stages of hypertension

The multivariable analysis presented in Table 2 reveals that male sex is significantly associated with higher odds of presenting all HTN stages, particularly in stages I and II. The results indicate that the odds of presenting HTN, regardless of the stage, also increase significantly as age increases.

**Table 2.**
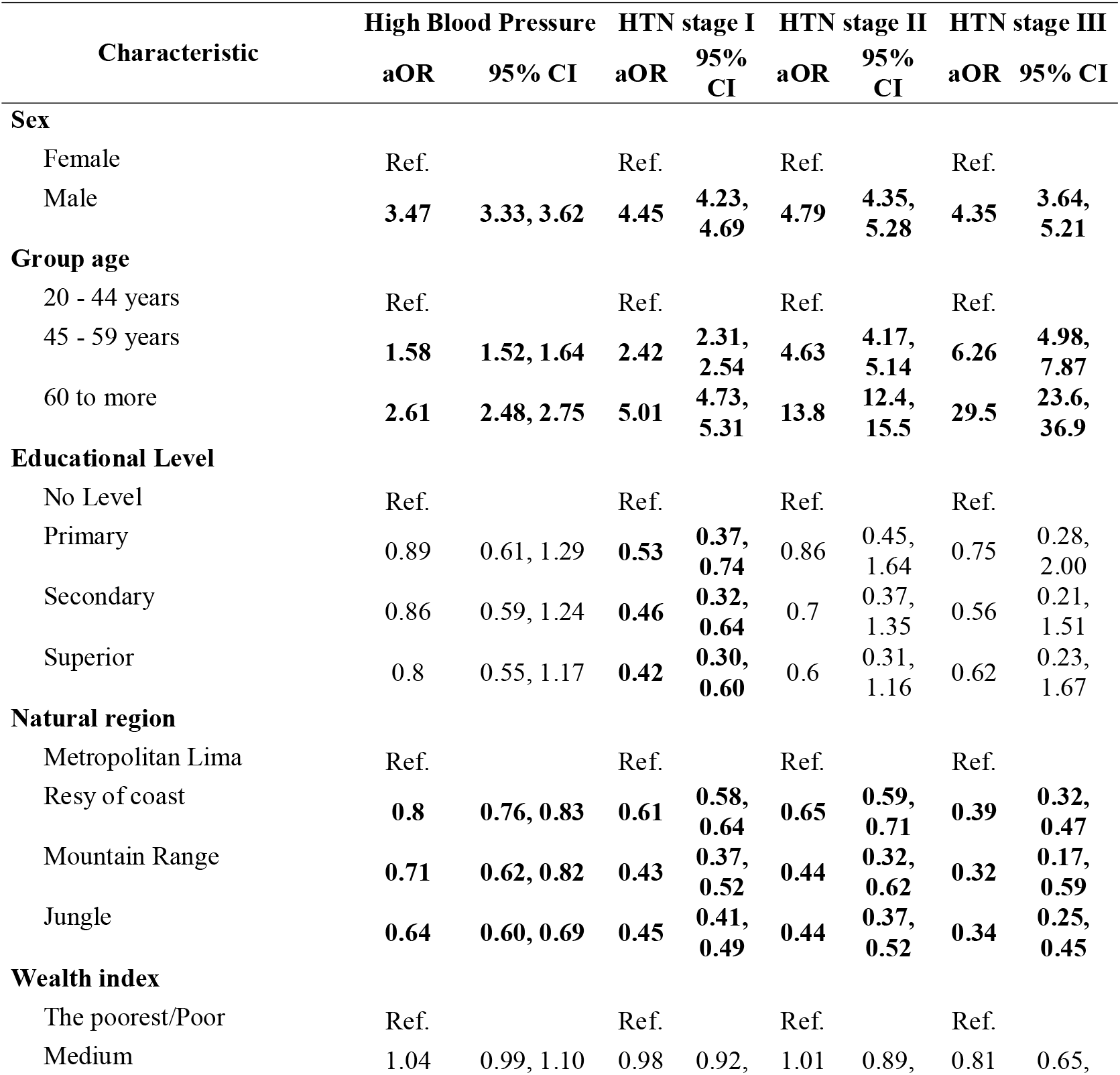
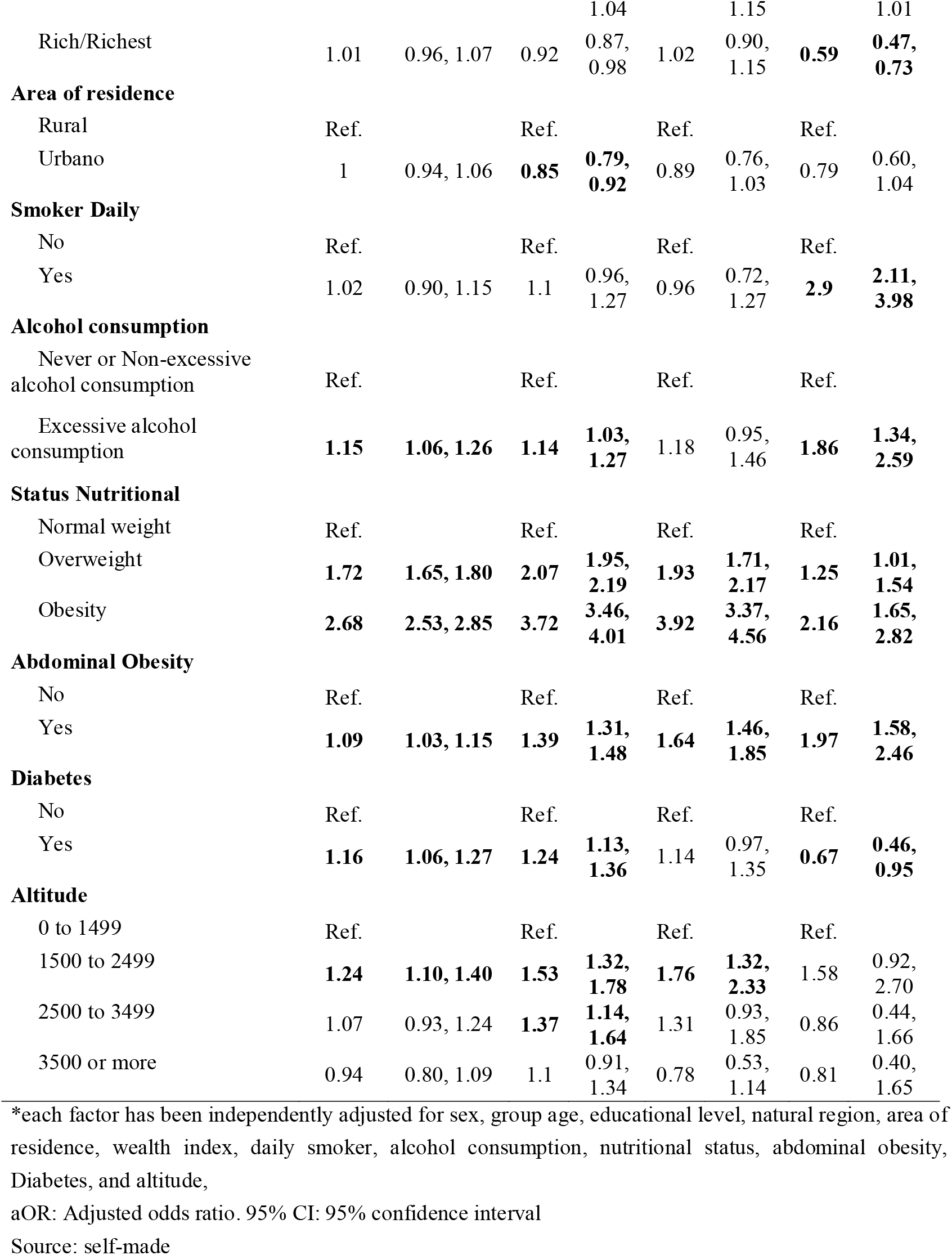
Multivariable regression analysis of the factors associated with the different types of HTN.

On the other hand, regions outside of Metropolitan Lima and rural areas show lower odds of all HTN stages than Metropolitan Lima. Regarding the location of residence, urban areas present a protective effect against stage I HTN.

In terms of lifestyle factors, smoking does not show a significant association with stage I or II but is associated with higher odds of stage III HTN. Excessive alcohol consumption is associated with increased odds of HBP, stage I HTN, and stage III HTN. In contrast, regular consumption of fruits and vegetables shows a protective effect against stage II HTN.

Regarding nutritional status, being overweight and obese significantly increases the odds of all stages of HTN, particularly obesity. Type 2 diabetes mellitus increases the odds of HBP and stage I HTN while showing a protective role against stage III HTN. Finally, altitude shows a varied pattern.

## Discussion

### Prevalence and trends of different stages of hypertension

The observation that the various stages of HTN do not show a clear upward trend but rather oscillate year by year represents a complex phenomenon reflecting multiple underlying influences, including population changes, public health interventions, and variations in treatment adherence. This fluctuation in HTN prevalence rates has been observed in several studies over time and can be attributed to social, economic, and health factors ^(11–15)^.

It is essential to note the prevalence of stage III HTN, also known in some international guidelines as hypertensive crisis. In our study, only 0.64% of the participants presented with this severe degree of hypertension. This figure is considerably lower compared to other international studies, where the prevalence tends to be higher. For instance, a recent systematic review that included eight observational studies in emergency departments reported a global prevalence of hypertensive crisis at 1.2%^(16)^, while the work of Bikbov et al. ^(11)^ reported a prevalence of 2.7%.

In the Peruvian population, the study by Calderón-Ocón et al. ^(6)^, conducted in 2024, found a hypertension prevalence of 1.5%. However, it is essential to note that this figure included both individuals aware of their hypertensive condition and those undiagnosed. Unlike this approach, our study chose to exclude individuals who were already aware of their HTN. This decision was based on the premise that those who know their hypertensive status may have taken measures to control their blood pressure, which could bias the estimation of the actual prevalence in the undiagnosed population. By focusing solely on individuals without a prior diagnosis, we aimed to obtain a clearer picture of the prevalence of different stages of hypertension in those who may be unaware of their condition. This methodology allows us to identify the need for intervention and awareness in the undiagnosed population more accurately.

### Factors associated with different stages of hypertension

The findings of this study, which demonstrate a higher prevalence of HTN in men than in women, are supported by multiple international studies. For example, various studies have found that men have an increased risk of developing hypertension compared to women in diverse regions ^(17–19)^. From a pathophysiological perspective, testosterone elevates blood pressure by increasing peripheral vascular resistance and fluid retention ^(20)^. On the other hand, it is essential to highlight the protective role of estrogens in premenopausal women, which reduces the vascular stress response and promotes vasodilation, potentially explaining the lower rates of hypertension in this group until menopause ^(21)^.

The progressive increase in HTN prevalence with age observed in our results, reaching even levels corresponding to stage III hypertension, reflects well-documented physiological and structural changes associated with aging. Previous studies have demonstrated that, with age, arteries undergo a hardening process known as arteriosclerosis, attributable to the accumulation of calcium deposits and changes in connective tissue, which increases vascular resistance and, consequently, blood pressure^(22,23)^. The declining renal function accompanying aging is also critical in raising blood pressure. The kidneys of older individuals show a reduced capacity to filter blood and eliminate excess sodium, thereby contributing to increased blood volume and blood pressure. This renal dysfunction is directly related to alterations in the renin-angiotensin-aldosterone system, a key regulator of electrolyte balance and blood pressure ^(24)^.

The observation that regions outside Metropolitan Lima show a reduced prevalence of HTN compared to the latter highlights potentially significant differences in environmental, socioeconomic, and lifestyle factors. Several studies have reported similar patterns in other metropolises, where densely populated urban areas tend to exhibit higher HTN rates. The protective effect observed in other regions may be related to diets maintaining higher consumption of fresh and less processed foods and a less sedentary lifestyle ^(25)^. For example, a systematic review found that urbanization was strongly associated with increased HTN prevalence, a phenomenon attributed to more sedentary lifestyles and high-sodium diets in urban areas ^(26)^. Additionally, urban areas like Metropolitan Lima are often associated with higher stress levels, environmental pollution, and more industrialized food options, known risk factors for this disease^(4,27)^.

The observed relationship between a higher wealth index and a lower prevalence of HTN, precisely stage III HTN, is consistent with the literature suggesting that better socioeconomic status can offer protection against more severe forms of HTN. This phenomenon has been explored in other studies, which indicate that individuals in higher economic strata tend to have more excellent education and, therefore, better health knowledge, promoting healthy lifestyles, including balanced diets and regular physical activity ^(28,29)^. These habits significantly contribute to the prevention of chronic conditions, including severe hypertension. Lower stress levels related to economic concerns and more excellent stability can positively influence cardiovascular health.

Regarding harmful habits, both excessive alcohol consumption and daily smoking are associated with HTN, precisely stage III in the latter case. Previous studies have demonstrated that these unhealthy habits are associated with an increased cardiovascular risk, including HTN ^(30)^. In the context of smoking, our results indicate that daily smoking is associated with a higher prevalence of advanced stages of HTN, more than with its initial levels. Various international studies have demonstrated the benefits of quitting smoking in reducing the risk of developing HTN ^(31,32)^; however, the literature also suggests that the cardiovascular damage related to smoking does not depend solely on whether a person smokes but also on the amount consumed, such as the number of packs per day ^(33,34)^. Therefore, while the harmful effects of tobacco are widely recognized, further research is needed to fully unravel these complexities and confirm whether the intensity of smoking habit differentially influences the development of HTN. This need for additional research does not diminish the certainty about the harmful effects of tobacco and the importance of avoiding its use.

Finally, the phenomenon where known diabetic patients present a higher prevalence of HBP and stage I HTN, but none with stage III, and instead a protective effect against stage III HTN, deserves detailed consideration. This pattern may be related to the more active management of their condition, which has been supported by studies suggesting that diabetic patients, aware of their disease, may be more committed to controlling their health, including blood pressure, to avoid severe complications^(35,36)^. Diabetic patients typically receive regular counseling on lifestyle changes and antihypertensive medications as part of their standard treatment, which may help control blood pressure levels before they reach more dangerous thresholds, such as those of stage III HTN.

### Contribution to the Field

Our work analyzes the overall prevalence of HTN. It delves into the assessment of the different stages of this condition, offering a more detailed perspective on how the disease is distributed among the population, unlike other studies that focus solely on HTN in general terms. This differentiation is crucial for public health as it allows policymakers and health professionals to understand better the variations in the severity of HTN and its evolution within specific groups, thereby facilitating the evaluation of the effectiveness of current interventions and the adaptation of management strategies.

Additionally, identifying trends in the different stages of hypertension can reveal population subgroups that face risks or are underserved by current health policies. This information is invaluable for directing interventions that effectively address observed disparities, such as socioeconomic, demographic, or regional differences, thereby promoting more significant health equity. For instance, strategies could include specialized programs for rural areas that face unique barriers to healthcare access or for urban populations that may be exposed to distinctive risk factors.

The detailed study of hypertension stages also supports strategic planning and resource optimization in the healthcare system, laying the groundwork for more effective management of future disease burdens. Moreover, it fosters innovation in prevention and treatment strategies, possibly including the development of new technologies or therapeutic approaches tailored to the needs of patients at different stages of hypertension. This approach improves cardiovascular health outcomes and contributes to economic sustainability by reducing costs associated with managing advanced complications of this disease.

### Limitations of the Study

One significant limitation of this study is its cross-sectional design, as it does not allow for establishing causal relationships between the studied variables. This means that we cannot definitively assert that one causes the other. Additionally, although the study utilized a large and diverse sample, the generalizability of the results may be affected by the quality and accuracy of self-reported data, especially regarding lifestyle habits and health conditions such as smoking and diabetes.

## Conclusions

The prevalence of HTN, depending on its stage, shows significant variations and notable fluctuations over time. Furthermore, factors such as sex, age, wealth index, and health conditions like obesity and diabetes significantly influence the prevalence of various stages of hypertension. These findings highlight the need for public health strategies that are personalized and differentiated according to the specific hypertension stage to be more effective.

Based on these findings, it is recommended that education and prevention programs be implemented tailored to the predominantly associated factors and the stage of HTN identified in different population subgroups. These interventions should aim to improve the detection and management of this disease and educate about the importance of early detection and continuous management of hypertension, regardless of the initially detected severity stage.

## Data Availability

The data supporting the findings of this study can be accessed at the following link: https://proyectos.inei.gob.pe/microdatos/

## Acknowledgments

We want to express my gratitude to the members of the Instituto de Investigación de Enfermedades Tropicales, Universidad Nacional Toribio Rodríguez de Mendoza de Amazonas (UNTRM), Amazonas, Perú, who provided valuable comments during the preparation of this study.

## Financial Disclosure

This study is self-financed.

## Conflict of Interest

The authors declare no conflict of interest.

## Informed Consent

It was not necessary to obtain informed consent in this study.

## Author Contributions

Fiorella E. Zuzunaga-Montoya: Conceptualization, Investigation, Methodology, Writing - Original Draft, Writing - Review & Editing.

Luisa Erika Milagros Vásquez-Romero: Investigation, Project Administration, Writing - Original Draft, Writing - Review & Editing.

Enrique Vigil-Ventura: Investigation, Resources, Writing - Original Draft, Writing - Review & Editing.

Joan A. Loayza-Castro: Software, Data Curation, Formal Analysis, Writing - Review & Editing.

Carmen Inés Gutierrez De Carrillo: Validation, Visualization, Writing - Original Draft, Writing - Review & Editing.

Víctor Juan Vera-Ponce: Methodology, Supervision, Funding Acquisition, Writing - Review & Editing.

## Notes

### Competing Interest Statement

The authors have declared no competing interest.

### Funding Statement

This study did not receive any funding

## References

1. Comparison of the ACC/AHA and ESC/ESH Hypertension Guidelines [Internet]. American College of Cardiology. [cited on March 7, 2024]. Available at: https://www.acc.org/Latest-in-Cardiology/Articles/2019/11/25/08/57/ www.acc.org%2fLatest-in-Cardiology%2fArticles%2f2019%2f11%2f25%2f08%2f57%2fComparison-of-the-ACC-AHA-and-ESC-ESH-Hypertension-Guidelines

2. The first WHO report details the devastating impact of hypertension and ways to stop it [Internet]. [cited on March 8, 2024]. Available at:: https://www.who.int/news/item/19-09-2023-first-who-report-details-devastating-impact-of-hypertension-and-ways-to-stop-it

3. World Health Organization. Hypertension [Internet]. 2023 Mar 16 [cited 2024 Mar 8]. Available from: https://www.who.int/news-room/fact-sheets/detail/hypertension

4. Vera-Ponce VJ, Zuzunaga-Montoya FE, Vásquez-Romero LEM, Loayza-Castro JA, Paucar CRI, Valladares-Garrido MJ, et al. Analysis of Hypertension in Peru: Prevalence, Associated Factors, Knowledge, Management and Control, 2014-2022. medRxiv; 2024. p. 2024.04.22.24306187. doi:10.1101/2024.04.22.24306187

5. Han M, Chen Q, Liu L, Li Q, Ren Y, Zhao Y, et al. Stage 1 hypertension by the 2017 American College of Cardiology/American Heart Association hypertension guidelines and risk of cardiovascular disease events: systematic review, meta-analysis, and estimation of population etiologic fraction of prospective cohort studies. J Hypertens. 2020;38(4):573. doi:10.1097/HJH.0000000000002321

6. Calderon-Ocon V, Cueva-Peredo F, Bernabe-Ortiz A. Prevalence, trends, and factors associated with hypertensive crisis among Peruvian adults. Cad Saude Publica. 2024;40(2):e00155123. doi:10.1590/0102-311XEN155123

7. PERÚ Instituto Nacional de Estadística e Informática [Internet]. [citado el 30 de noviembre de 2021]. Disponible en: http://iinei.inei.gob.pe/microdatos/

8. Von Elm E, G.Altman D, Egger M, J.Pocock S, C.Gotzsche P, P.Vandenbrouckef J. Declaración de la Iniciativa STROBE (Strengthening the Reporting of Observational studies in Epidemiology): directrices para la comunicación de estudios observacionales. 22(2):144–50. doi:https://www.equator-network.org/wp-content/uploads/2015/10/STROBE_Spanish.pdf

9. NCD Risk Factor Collaboration (NCD-RisC). Worldwide trends in blood pressure from 1975 to 2015: a pooled analysis of 1479 population-based measurement studies with 19·1 million participants. Lancet Lond Engl. 2017;389(10064):37–55. doi:10.1016/S0140-6736(16)31919-5

10. PERU MIGRANT Study | Baseline dataset [Internet]. figshare; 2016 [cited on March 14, 2021]. doi:10.6084/m9.figshare.3125005.v1

11. Bikbov MM, Kazakbaeva GM, Zainullin RM, Salavatova VF, Gilmanshin TR, Yakupova DF, et al. Prevalence, Awareness, and Control of Arterial Hypertension in a Russian Population. The Ural Eye and Medical Study. Front Public Health [Internet]. 2020 [citado el 13 de mayo de 2024];7. doi:10.3389/fpubh.2019.00394

12. Wu J, Duan W, Jiao Y, Liu S, Zheng L, Sun Y, et al. The Association of Stage 1 Hypertension, Defined by the 2017 ACC/AHA Guidelines, With Cardiovascular Events Among Rural Women in Liaoning Province, China. Front Cardiovasc Med. 2021;8:710500. doi:10.3389/fcvm.2021.710500

13. Ostchega Y, Hughes JP, Zhang G, Nwankwo T, Graber J, Nguyen DT. Differences in Hypertension Prevalence and Hypertension Control by Urbanization Among Adults in the United States, 2013-2018. Am J Hypertens. 2022;35(1):31–41. doi:10.1093/ajh/hpab067

14. Ostchega Y, Hughes JP, Kit B, Chen T-C, Nwankwo T, Commodore-Mensah Y, et al. Differences in Hypertension and Stage II Hypertension by Demographic and Risk Factors, Obtained by Two Different Protocols in US Adults: National Health and Nutrition Examination Survey, 2017-2018. Am J Hypertens. 2022;35(7):619–26. doi:10.1093/ajh/hpac042

15. Mumena WA, Hammouda SA, Aljohani RM, Alzahrani AM, Bamagos MJ, Alharbi WK, et al. Prevalence and determinants of undiagnosed hypertension in the Western region of Saudi Arabia. PloS One. 2023;18(3):e0280844. doi:10.1371/journal.pone.0280844

16. Astarita A, Covella M, Vallelonga F, Cesareo M, Totaro S, Ventre L, et al. Hypertensive emergencies and urgencies in emergency departments: a systematic review and meta-analysis. J Hypertens. 2020;38(7):1203–10. doi:10.1097/HJH.0000000000002372

17. Redfern A, Peters SAE, Luo R, Cheng Y, Li C, Wang J, et al. Sex differences in the awareness, treatment, and control of hypertension in China: a systematic review with meta-analyses. Hypertens Res Off J Jpn Soc Hypertens. 2019;42(2):273–83. doi:10.1038/s41440-018-0154-x

18. Joo HJ, Yum Y, Kim YH, Son J-W, Kim SH, Choi S, et al. Gender Difference of Blood Pressure Control Rate and Clinical Prognosis in Patients With Resistant Hypertension: Real-World Observation Study. J Korean Med Sci. 2023;38(16):e124. doi:10.3346/jkms.2023.38.e124

19. Connelly PJ, Currie G, Delles C. Sex Differences in the Prevalence, Outcomes and Management of Hypertension. Curr Hypertens Rep. 2022;24(6):185–92. doi:10.1007/s11906-022-01183-8

20. Gerdts E, Sudano I, Brouwers S, Borghi C, Bruno RM, Ceconi C, et al. Sex differences in arterial hypertension. Eur Heart J. 2022;43(46):4777–88. doi:10.1093/eurheartj/ehac470

21. Mendelsohn ME, Karas RH. The protective effects of estrogen on the cardiovascular system. N Engl J Med. 1999;340(23):1801–11. doi:10.1056/NEJM199906103402306

22. Nilsson PM. Early Vascular Aging in Hypertension. Front Cardiovasc Med [Internet]. 2020 [citado el 13 de mayo de 2024];7. doi:10.3389/fcvm.2020.00006

23. Strandberg TE. Blood Pressure in a 100-Year Perspective. Circulation. 2019;140(2):101–2. doi:10.1161/CIRCULATIONAHA.119.040168

24. Glassock RJ, Rule AD. Aging and the Kidneys: Anatomy, Physiology and Consequences for Defining Chronic Kidney Disease. Nephron. 2016;134(1):25–9. doi:10.1159/000445450

25. Bernabé-Ortiz A, Carrillo-Larco RM, Gilman RH, Checkley W, Smeeth L, Miranda JJ. Impact of urbanisation and altitude on the incidence of, and risk factors for, hypertension. Heart. 2017;103(11):827–33. doi:10.1136/heartjnl-2016-310347

26. Mills KT, Bundy JD, Kelly TN, Reed JE, Kearney PM, Reynolds K, et al. Global Disparities of Hypertension Prevalence and Control: A Systematic Analysis of Population-based Studies from 90 Countries. Circulation. 2016;134(6):441–50. doi:10.1161/CIRCULATIONAHA.115.018912

27. Wu J, Chen J, Li Z, Jiao B, Muennig P. Spatiotemporal Variation of the Association between Urbanicity and Incident Hypertension among Chinese Adults. Int J Environ Res Public Health. 2020;17(1):304. doi:10.3390/ijerph17010304

28. Steptoe A, Marmot M. The role of psychobiological pathways in socio-economic inequalities in cardiovascular disease risk. Eur Heart J. 2002;23(1):13–25. doi:10.1053/euhj.2001.2611

29. Cerpa-Arana SK, Rimarachín-Palacios LM, Bernabé-Ortiz A. Association between socioeconomic level and cardiovascular risk in the Peruvian population. Rev Saude Publica. 2022;56:91. doi:10.11606/s1518-8787.2022056004132

30. Unger T, Borghi C, Charchar F, Khan NA, Poulter NR, Prabhakaran D, et al. 2020 International Society of Hypertension Global Hypertension Practice Guidelines. Hypertension. 2020;75(6):1334–57. doi:10.1161/HYPERTENSIONAHA.120.15026

31. Datta BK, Husain MJ. Uncontrolled hypertension among tobacco-users: women of prime childbearing age at risk in India. BMC Womens Health. 2021;21(1):146. doi:10.1186/s12905-021-01280-x

32. Lynch J, Jin L, Richardson A, Conklin DJ. Tobacco Smoke and Endothelial Dysfunction: Role of Aldehydes? Curr Hypertens Rep. 2020;22(9):73. doi:10.1007/s11906-020-01085-7

33. Kondo T, Nakano Y, Adachi S, Murohara T. Effects of Tobacco Smoking on Cardiovascular Disease. Circ J Off J Jpn Circ Soc. 2019;83(10):1980–5. doi:10.1253/circj.CJ-19-0323

34. Kim SJ, Jee SH, Nam JM, Cho WH, Kim J-H, Park E-C. Do early onset and pack-years of smoking increase risk of type II diabetes? BMC Public Health. 2014;14:178. doi:10.1186/1471-2458-14-178

35. Volpe M, Battistoni A, Savoia C, Tocci G. Understanding and treating hypertension in diabetic populations. Cardiovasc Diagn Ther. 2015;5(5):353–63. doi:10.3978/j.issn.2223-3652.2015.06.02

36. Lara-Esqueda A, Zaizar-Fregoso SA, Madrigal-Perez VM, Ramirez-Flores M, Montes-Galindo DA, Martinez-Fierro ML, et al. Evaluation of Medical Care for Diabetic and Hypertensive Patients in Primary Care in Mexico: Observational Retrospective Study. J Diabetes Res. 2021;2021:7365075. doi:10.1155/2021/7365075

